# A brief analysis of the COVID-19 death data in Malaysia

**DOI:** 10.1101/2021.09.28.21264234

**Authors:** Wan Nor Arifin, Kamarul Imran Musa, Tengku Muhammad Hanis, Wan Shakira Rodzlan Hasani, Che Muhammad Nur Hidayat Che Nawi, Erwan Ershad Ahmad Khan, Mohd Azmi Suliman, Sahrol Azmi Termizi, Wira Alfatah Ab Aziz, on behalf of the USM Epidemiological Modelling Team

## Abstract

In December 2019, the first cases of Coronavirus Disease 2019 (COVID-19) were detected in Wuhan, China. Since then, COVID-19 begun to spread rapidly all over the world. On March 11, 2020, the World Health Organisation declared COVID-19 a pandemic. As of September 7, 2021, there were over 220 million confirmed COVID-19 cases globally, with more than 4.6 million deaths. Malaysia reported 2,067,327 confirmed cases with 22,743 deaths. Given the severity of the pandemic, the Ministry of Health Malaysia has stepped up in its efforts in handling the pandemic locally by sharing the COVID-19 related data on the GitHub, enabling transparent data sharing. This enables timely data analysis and quick decision to better understand the COVID-19 situation in this country. This article aims to provide a quick analysis of the death and vaccination data provided by the Malaysian Ministry of Health and to provide useful insight into the analysis.

## 1 Introduction

In December 2019, the first penumonia cases of unknown origin - later identified Coronavirus Disease 2019 (COVID-19) - were detected in Wuhan, China. Since then, it begun to spread rapidly across mainland China and all over the world. On March 11, 2020, the World Health Organisation declared COVID-19 as a pandemic (1). As of September 7, 2021, COVID-19 has infected over 220 million (226,844,344) people worldwide with more than 4.6 million (4,666,334) deaths (2). On the same day, Malaysia reported 2,067,327 confirmed cases with 22,743 deaths (3). Numerous studies have shown that deaths related to COVID-19 have been linked to elderly, male, and various pre-existing conditions such as cardiovascular disease, diabetes, hypertension, respiratory disease, and hypertension (4–7).

Different countries have formulated different containment and mitigation strategies to combat this disease, including lockdowns and curfews while anxiously waiting for the arrival of definitive tool to fight this disease. Vaccine development was accelerated to achieve immunity to the virus and prevent transmission (8). By mid-2021, 3 billion doses of the COVID-19 vaccine had been administered worldwide, with more than 40 million doses of the COVID-19 vaccine administered worldwide every day (9). In Malaysia, vaccination coverage was selected as one of the key threshold indicators for the National Recovery Plan for COVID-19 pandemic (10).

On February 24, 2021, Malaysia launched its vaccination campaign against COVID-19. In the early phases, the campaign prioritised the at risk population; namely the healthcare workers and elderly with comorbidities after the approval for the use of the COVID-19 vaccines, Cominarty® (Pfizer-BioNTech) and CoronaVac® (Sinovac). Currently, the number of COVID-19 vaccines available in Malaysia has increased to five, including ChAdOx1-S (Oxford-AstraZeneca), Ad26.COV2-S® (Janssen) and Convidecia(TM) (CanSinoBio) (10). By September 17, 2021, around 55.5% of the total population has completed the vaccination and at least 66.6% of them have completed the first dose of vaccination (11,12).

The vaccination campaign has the potential to reduce the burden of COVID-19 in terms of morbidity and mortality if high vaccine intake is achieved. A previous study reported that the risk of COVID-19 infection, hospitalization and death was significantly reduced in vaccinated people (13). However, the effectiveness of vaccination in lowering the death rate may vary depending on the type of COVID-19 vaccine.

With COVID-19 affecting mortality rapidly around the world, understanding and assessing the risks of death from COVID-19 infection through timely data analysis is critical. Traditional approaches previously used in research, such as extracting data from the electronic health record (HER), resulted in late reports resulting in delayed action. The timeliness in term of initial diagnosis to record, and record to report may vary from 6 months to 4 years respectively (14). This is due to decentralized data management and sharing within and between ministries and other organizations. A novel approach by the Ministry of Health Malaysia by sharing the COVID-19 related data on the GitHub has enabled transparent data sharing. This will allow timely data analysis and quick decision to better understand the COVID-19 situation in this country. This article aims to provide a quick analysis of the death and vaccination data provided by the Malaysian Ministry of Health (11,15) and to provide useful insight into the analysis.

## 2 Method

### 2.1 Data

The data was sourced from MoH Malaysia’s GitHub repository at https://github.com/MoH-Malaysia/covid19-public/tree/main/epidemic/linelist as of Sep 28, 2021 (15). The `linelist_deaths.csv` contains:

In addition, vaccination data was sourced from Malaysia’s National COVID-19 Immunisation Programme (or *Program Imunisasi COVID-19 Kebangsaan*, PICK)’s GitHub repository at https://github.com/CITF-Malaysia/citf-public as of Sep 28, 2021 (11). `The vax_malaysia.csv` contains:

The death cases were classified as:

1. `Unvaccinated`: Cases who had not received any COVID-19 vaccination.
2. `Partial`: Partially vaccinated when the cases were tested COVID-19 positive (< 14 days after the last dose of vaccination).
3. `Complete`: Completed vaccination when the cases were tested COVID-19 positive (> 14 days after the last dose of vaccination).

### 2.2 Statistical Analysis

R version 3.6.3 was used. In general, `tidyverse` (16), `gtsummary` (17), `broom` (18) and `knitr` (19) R packages were used. We used quasipoisson regression to analyze the count data for deaths in all groups. Next, logistic regression with frequency weights were utilized, where we combined the death data with the vaccination data to utilize the number of completely vaccinated individuals in the model. Updated document in HTML format and detailed data management and analysis are available at https://healthdata.usm.my:3939/content/248.

## 3 Results and Insights

### 3.1 Analysis for all deaths

The descriptive analysis of deaths among all reported cases (N = 25,935) is presented in Table 3. Unvaccinated group reported the highest percentage of death with 18,133 (69.9%), followed by partially vaccinated and completely vaccinated groups with 5,846 (22.5%) and 1,956 (7.5%), respectively. The mean age for unvaccinated and partially vaccinated groups was lower than the completely vaccinated group, indicating more cases at relatively a younger age died in unvaccinated and partially vaccinated groups. The percentage with comorbid conditions for unvaccinated and partially vaccinated groups was lower than the completely vaccinated group, indicating more people without comorbid conditions died in unvaccinated and partially vaccinated groups.

**Table 1:**
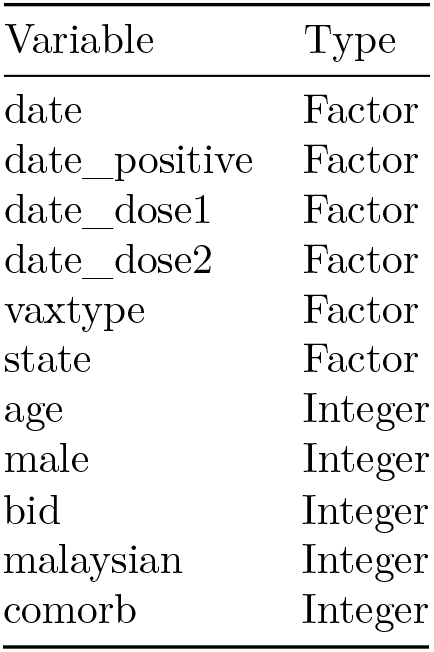
Variable names and types of death data.

| Variable | Type |
| --- | --- |
| date | Factor |
| date__positive | Factor |
| date__dose1 | Factor |
| date__dose2 | Factor |
| vaxtype | Factor |
| state | Factor |
| age | Integer |
| male | Integer |
| bid | Integer |
| malaysian | Integer |
| comorb | Integer |

**Table 2:**
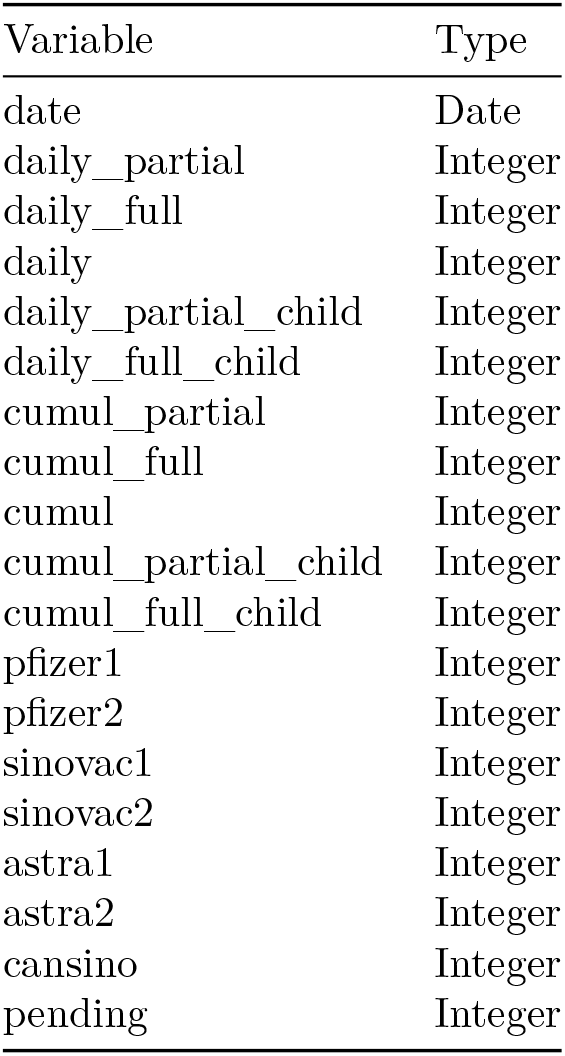
Variable names and types of vaccination data.

**Table 3:** Descriptive analysis for all deaths.

| Characteristic | Complete, N =<br>1,956 | Partial, N = 5,846 | Unvaccinated, N =<br>18,133 |
| --- | --- | --- | --- |
| **Vaccination Type** |  |  |  |
| AstraZeneca | 18 (0.9%) | 697 (12%) | 0 (0%) |
| Pending VMS sync | 1 (<0.1%) | 25 (0.4%) | 0 (0%) |
| Pfizer | 439 (22%) | 2,008 (34%) | 0 (0%) |
| Sinovac | 1,498 (77%) | 3,116 (53%) | 0 (0%) |
| No Vaccine | 0 (0%) | 0 (0%) | 18,133 (100%) |
| **Age** | 67.3 (13.1) | 59.8 (14.5) | 59.8 (16.5) |
| **Gender (Male)** | 1,177 (60%) | 3,415 (58%) | 10,282 (57%) |
| **BID** | 378 (19%) | 930 (16%) | 3,860 (21%) |
| **Malaysian** | 1,845 (94%) | 5,421 (93%) | 15,219 (84%) |
| **Comorbidity** | 1,645 (84%) | 4,362 (75%) | 13,295 (73%) |
| **Test Positive to Death**<br>**(days)** | 5.3 (6.3) | 6.9 (9.5) | 6.8 (8.9) |
| **State** |  |  |  |
| Johor | 302 (15%) | 767 (13%) | 2,172 (12%) |
| Kedah | 136 (7.0%) | 388 (6.6%) | 1,301 (7.2%) |
| Kelantan | 64 (3.3%) | 129 (2.2%) | 604 (3.3%) |
| Melaka | 64 (3.3%) | 223 (3.8%) | 563 (3.1%) |
| Negeri Sembilan | 52 (2.7%) | 244 (4.2%) | 897 (4.9%) |
| Pahang | 46 (2.4%) | 96 (1.6%) | 440 (2.4%) |
| Perak | 103 (5.3%) | 177 (3.0%) | 549 (3.0%) |
| Perlis | 3 (0.2%) | 17 (0.3%) | 42 (0.2%) |
| Pulau Pinang | 191 (9.8%) | 273 (4.7%) | 851 (4.7%) |
| Sabah | 170 (8.7%) | 294 (5.0%) | 1,715 (9.5%) |
| Sarawak | 153 (7.8%) | 86 (1.5%) | 555 (3.1%) |
| Selangor | 457 (23%) | 2,441 (42%) | 6,365 (35%) |
| Terengganu | 30 (1.5%) | 72 (1.2%) | 257 (1.4%) |
| W.P. Kuala Lumpur | 184 (9.4%) | 621 (11%) | 1,671 (9.2%) |
| W.P. Labuan | 1 (<0.1%) | 12 (0.2%) | 135 (0.7%) |
| W.P. Putrajaya | 0 (0%) | 6 (0.1%) | 16 (<0.1%) |

To determine the risk factors with death due to COVID-19, quasi-poisson regression was performed in view of the presence of overdispersion in the data. The results are displayed in Table 4. Partially and completely vaccinated groups showed 4.9 and 8.8 times lower risk of death as compared to unvaccinated group (IRR < 1 indicates protective effect, where x times lower risk equals 1/IRR). Other risk factors, where were age of 60 and above, male gender, and having commorbid conditions were at higher risk of death at 1.3, 1.2 and 2.6 times higher risk of death.

**Table 4:**
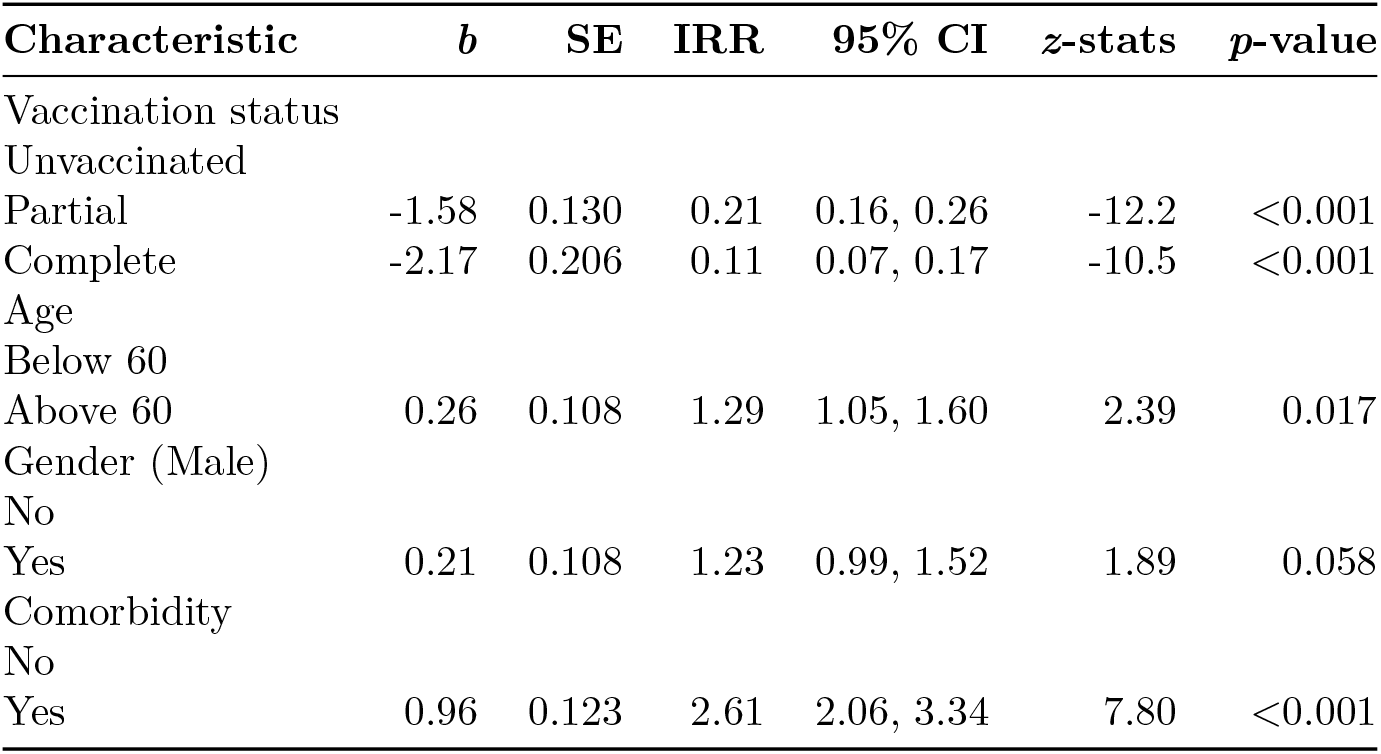
Risk factors of death by quasipoisson regression model, (N = 25,909).

### 3.2 Analysis for completely vaccinated deaths

The descriptive analysis of deaths among the completely vaccinated cases (N = 1,956) is presented in Table 5. Sinovac group reported the highest percentage of in-group death with 1,498 (76.6%), followed by Pfizer group with 439 (22.5%). AstraZeneca group reported the smallest number of deaths with 18 (0.9%) (see also Table 3, Column 1, Complete). This descriptive analysis is followed by a detailed sub-group analysis.

**Table 5:**
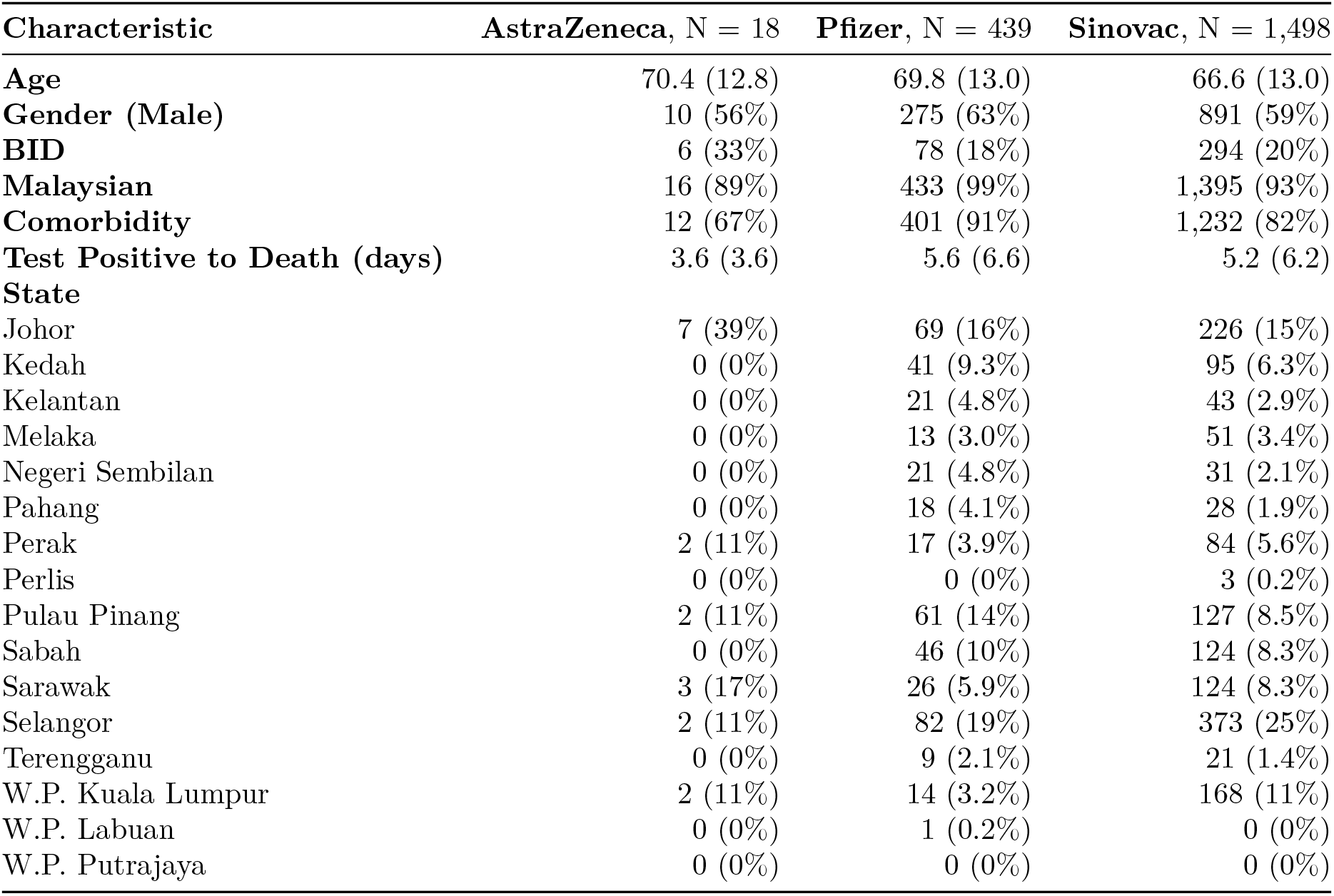
Descriptive analysis for completely vaccinated deaths.

In this sub-group analysis, stratified death rates are calculated by vaccine type (Table 6). We took the number of vaccinated individuals at 14 days earlier from the date of latest death among completely vaccinated individuals. This was done to ensure that death in the data analysed occurred only after the cases completed the vaccination 14 days earlier.

**Table 6:** Sub-group analysis for completely vaccinated deaths.

| Vaccine Type | Completed Vaccination | Death | Death Rate per 100,000 |
| --- | --- | --- | --- |
| AstraZeneca | 1,072,083 (6.11%) | 18 (0.92%) | 1.68 |
| Pfizer | 7,589,773 (43.24%) | 439 (22.46%) | 5.78 |
| Sinovac | 8,702,598 (49.58%) | 1,498 (76.62%) | 17.21 |

Here, death rate is defined as:

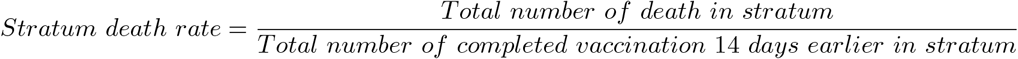

where, in relation to Table 2, the total number of completed vaccination per vaccine type was calculated from `astra2`, pfizer2, `and`sinovac2’ variables.

The death rate per 100,000 individuals is notably higher in the Sinovac group, followed by the Pfizer group. To determine the difference in death rate across the groups is significant statistically, we performed a weighted logistic regression, where we combined the death data with the vaccination data to utilize the number of completely vaccinated individuals in the model. The logistic regression was performed to determine whether vaccine type is associated with death. Again, we reiterate that we took the number of vaccinated individuals at 14 days earlier from the date of latest death among completely vaccinated individuals to ensure that death in the data analysed occurred only after the cases completed the vaccination 14 days earlier. The weighted logistic regression is presented in Table 7.

**Table 7:** Association between vaccine type and death among completely vaccinated persons, (N = 17,364,454).

| Characteristic | *b* | SE | OR | 95% CI | *z*-stats | *p*-value |
| --- | --- | --- | --- | --- | --- | --- |
| Vaccine Type |  |  |  |  |  |  |
| AstraZeneca |  |  |  |  |  |  |
| Pfizer | 1.24 | 0.240 | 3.45 | 2.22, 5.73 | 5.14 | <0.001 |
| Sinovac | 2.33 | 0.237 | 10.3 | 6.66, 17.0 | 9.82 | <0.001 |

The odds ratios for Pfizer vs AstraZeneca and Sinovac vs AstraZeneca are OR = 3.45 (95% CI: 2.22, 5.73) and OR = 10.25 (95% CI: 6.66, 16.95) respectively, of which both odds ratios are statistically significant (Table 7). These odds ratios indicate that Pfizer and Sinovac groups had higher odds of death as compared to AstraZeneca group. However, we suspected that this finding could be biased, because in the AztraZeneca group, the time between the first and second dose is typically eight weeks and above, so some deaths might have happened in between this period (see Table 3, Column 2, Partial). So, we proceeded with another analysis which combined partially and completely vaccinated groups in the following section.

### 3.3 Analysis for partially and completely vaccinated deaths

The descriptive analysis of deaths among the partially and completely vaccinated cases (N = 7,802) is presented in Table 8. Sinovac group reported the highest percentage of in-group death with 4614 (59.3%), followed by Pfizer group with 2447 (31.5%). AstraZeneca group reported the smallest number of deaths with 715 (9.2%) (see also Table 3, Column 1, Complete). This descriptive analysis is followed by a detailed sub-group analysis.

**Table 8:** Descriptive analysis for partially and completely vaccinated deaths.

| Characteristic | AstraZeneca, N = 715 | Pfizer, N = 2,447 | Sinovac, N = 4,614 |
| --- | --- | --- | --- |
| Age | 64.0 (14.7) | 62.3 (14.7) | 61.0 (14.3) |
| Gender (Male) | 432 (60%) | 1,409 (58%) | 2,739 (59%) |
| BID | 81 (11%) | 399 (16%) | 817 (18%) |
| Malaysian | 684 (96%) | 2,356 (96%) | 4,204 (91%) |
| Comorbidity | 545 (76%) | 2,000 (82%) | 3,445 (75%) |
| Test Positive to Death (days) | 7.2 (7.8) | 6.4 (9.4) | 6.4 (8.7) |
| State |  |  |  |
| Johor | 127 (18%) | 395 (16%) | 545 (12%) |
| Kedah | 32 (4.5%) | 261 (11%) | 229 (5.0%) |
| Kelantan | 6 (0.8%) | 89 (3.6%) | 95 (2.1%) |
| Melaka | 14 (2.0%) | 87 (3.6%) | 186 (4.0%) |
| Negeri Sembilan | 6 (0.8%) | 136 (5.6%) | 154 (3.3%) |
| Pahang | 2 (0.3%) | 70 (2.9%) | 70 (1.5%) |
| Perak | 16 (2.2%) | 90 (3.7%) | 172 (3.7%) |
| Perlis | 0 (0%) | 8 (0.3%) | 12 (0.3%) |
| Pulau Pinang | 37 (5.2%) | 191 (7.8%) | 234 (5.1%) |
| Sabah | 21 (2.9%) | 210 (8.6%) | 227 (4.9%) |
| Sarawak | 3 (0.4%) | 50 (2.0%) | 185 (4.0%) |
| Selangor | 353 (49%) | 697 (28%) | 1,843 (40%) |
| Terengganu | 1 (0.1%) | 36 (1.5%) | 65 (1.4%) |
| W.P. Kuala Lumpur | 97 (14%) | 110 (4.5%) | 595 (13%) |
| W.P. Labuan | 0 (0%) | 12 (0.5%) | 1 (<0.1%) |
| W.P. Putrajaya | 0 (0%) | 5 (0.2%) | 1 (<0.1%) |

In this sub-group analysis, stratified death rates are calculated by vaccine type (Table 9). We took the latest number of vaccinated individuals as the denominator, because we no longer consider the completion of second dose as a criteria for the analysis. Here, death rate is defined as:

**Table 9:**
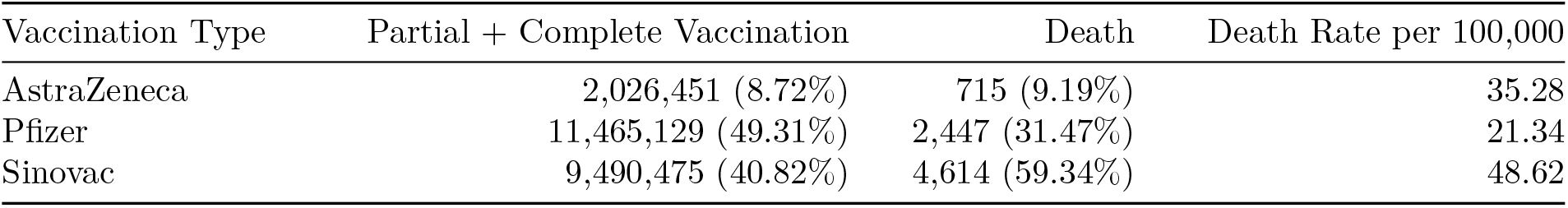
Stratified analysis for partially and completely vaccinated deaths.

| Vaccination Type | Partial + Complete Vaccination | Death | Death Rate per 100,000 |
| --- | --- | --- | --- |
| AstraZeneca | 2,026,451 (8.72%) | 715 (9.19%) | 35.28 |
| Pfizer | 11,465,129 (49.31%) | 2,447 (31.47%) | 21.34 |
| Sinovac | 9,490,475 (40.82%) | 4,614 (59.34%) | 48.62 |

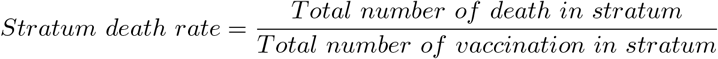

where, in relation to Table 2, the total number of vaccination per vaccine type was calculated from `astra1`, `pfizer1`, and `sinovac1` variables.

Table 9 shows a different picture to that of Table 6. The death rate per 100,000 individuals is highest in the AstraZaneca group, followed by the Sinovac group. Again, to determine the difference in death rate across the groups is significant statistically, we performed a weighted logistic regression, where we combined the death data with the vaccination data to utilize the number of vaccinated individuals in the model. The logistic regression was performed to determine whether vaccine type is associated with death. The weighted logistic regression is presented in Table 10.

**Table 10:** Association between vaccine type and death among partially and completely vaccinated persons, (N = 22,982,055).

| Characteristic | *b* | SE | OR | 95% CI | *z*-stats | *p*-value |
| --- | --- | --- | --- | --- | --- | --- |
| Vaccine Type |  |  |  |  |  |  |
| AstraZeneca |  |  |  |  |  |  |
| Pfizer | -0.50 | 0.043 | 0.60 | 0.56, 0.66 | -11.8 | <0.001 |
| Sinovac | 0.32 | 0.040 | 1.38 | 1.27, 1.49 | 7.98 | <0.001 |

The odds ratios for Pfizer vs AstraZeneca and Sinovac vs AstraZeneca are OR = 0.605 (95% CI: 0.557, 0.658) and OR = 1.378 (95% CI: 1.275, 1.492) respectively, of which both odds ratios are statistically significant (Table 10). These odds ratios indicate that Pfizer group had slightly lower odds of death (1.7 times lower) as compared to AstraZeneca group, while Sinovac group had slightly higher odds of death (1.4 times higher) as compared to AstraZeneca group. The advantage of Astrazeneca over the other two vaccines was diluted whenever we also included partially vaccinated individuals into analysis.

### 3.4 Summary of findings

Based on these analyses, we concluded that:

1. Vaccination, either partial or complete, tremendously reduced the risk of death by 4.9 and 8.8 times respectively (Table 4). This was based on the analysis for all COVID-19 deaths (Section 3.1).
2. Vaccine type was found to be associated with the odds of death, where Pfizer and Sinovac had higher odds of death (3.5 and 10.2 odds ratio respectively) as compared to AstraZeneca (Table 7). This was based on the analysis for completely vaccinated individuals (Section 3.2). However, we noted this could be attributed to the longer gap between first and second dose for AstraZeneca as compared to the other two vaccines.
3. Vaccine type was again found to be associated with the odds of death (Table 10). However, in contrast to the result in (2), Pfizer and Sinovac instead showed 0.6 and 1.4 odds ratios respectively when the analysis was based on the data for both partially and completely vaccinated individuals (Section 3.3). This result indicates that Pfizer group had slightly lower odds of death (1.7 times lower), while Sinovac group had slightly higher odds of death (1.4 times higher) in comparison to AstraZeneca group. However, we noted the differences between the vaccine types were quite small in magnitude despite their statistical significance.

### 3.5 Limitations

As of writing, there were several issues that limited our analyses. First, there was no line listing or per observation data for each vaccinated individuals. Although we had basic demographic information for the vaccinated death cases, the absence of information for the control group (vaccinated, alive) limited the logistic regression analysis to only the type of vaccine taken. Second, currently there was no line listing for each COVID-19 cases. The availability of this data will allow more insights into the associated factors of COVID-19 death.

## 4 Conclusion and Recommendations

Based on the findings, we clearly highlighted the advantage of COVID-19 vaccination in reducing the risk of death. Although there were advantages of choosing certain vaccines over others, these advantages negated each other when we took into account partial or complete vaccination status. In this respect, regardless of the vaccine type, getting vaccinated is the best way to protect against the risk of COVID-19 death.

## Data Availability

The data used in the article are publicly available.

https://github.com/MoH-Malaysia/covid19-public

https://github.com/CITF-Malaysia/citf-public

## Acknowledgement

We are grateful to the Ministry of Health of Malaysia (MoH) for their transparent data sharing initiative. This analysis would be not be possible without this data sharing initiative by the MoH. We also thank Dr. Shahnon Anuar Shahrani for proofreading the draft of this preprint.

## USM Epidemiological Modelling Team

Department of Community Medicine, School of Medical Sciences, Universiti Sains Malaysia:

- Assoc. Prof. Dr. Kamarul Imran Musa
- Tengku Muhammad Hanis Tengku Mokhtar
- Dr. Mohd Azmi Suliman
- Dr. Che Muhammad Nur Hidayat Che Nawi

Biostatistics and Research Methodology Unit, School of Medical Sciences, Universiti Sains Malaysia:

- Dr. Wan Nor Arifin

Faculty of Medicine, Universiti Teknologi MARA,

- Dr. Xin Wee Chen

Ministry of Health, Malaysia:

- Dr. Afiqah Syamimi Masrani
- Dr. Erwan Ershad Ahmad Khan
- Dr. Mohamad Zarudin Mat Said
- Dr. Sahrol Azmi Termizi
- Dr. Wira Alfatah Ab Ayah @ Ab Aziz
- Wan Shakira Rodzlan Hasani

Pusat Data PPKT, Health Campus, Universiti Sains Malaysia:

- Mohd Fadzali Bakar
- Mohd Faizal Abdul Manaf
- Ahmad Syakiren Mazalan

